## Supplementary material for "Clinical, immune and genetic risk factors of malaria-associated acute kidney injury in Zambian Children: A study protocol": S1 effect sizes

**Sub-study 1 sample size considerations**

Objective 1**:** The sample size was determined using computer software. For categorical variables OpenEPI which uses the formula for two proportions was utilized, while for continuous variables the power calculation function for independent t-test for two samples in SPSS version 29 was used. Non-parametric data was adjusted to enable use of this method as described elsewhere. To detect an effect size (proportions for categorical variables and magnitudes for continuous variables), findings from previous studies were used. Other assumptions made were a statistical significance of 5%, a power of 80% and a case-to -control ratio of 1:1. Using this method, the sample size was chosen as shown in **S1 Table 1** with 190 children with MAKI (cases) to be recruited and 190 controls to be recruited giving a total sample size of 380.

**S1 Table 1:sample size estimation for objective 1 (power 80%, statistical significance 5%)**

| Clinical  Variable | N | Proportion/mean of  Controls with exposure | Proportion/mean/median  Of  Cases  With  Exposure | Estimated  Sample  Size |
| --- | --- | --- | --- | --- |
| Dehydration:  BUN: Cr ratio >20 [1] | 178 | 8% | 84% | 16 |
| Blackwater fever [2] | 598 | 17.2% | 31.5% | 186 |
| Hyperbilirubinemia  (mg/dl) [3] | 598 | 0·5(0·2, 0·9) | 0·9 (0·5, 1·7) | 110 |
| Mean serum haemoglobin [4] | 178 | 4.8 (3.3, 6.5) | 4. (4.0, 6.7) | 242 |
| Oxidative stress  MDA levels ( (µmol l−1) [5] | 100 | 0.47 ± 0.07 | 0.72 ± 0.25 | 22 |

Objective 2: For the immune molecule study, IBM TM-SPSS TM version 29 (IBM Corp., Armonk, NY, USA) was used to conduct power calculations based on previous studies that analysed cytokine/adhesion molecule association to severe malaria. **S2 Table 2** shows these studies. Based on this, 200 malaria patients (100 cases and 100 controls) will be used. This is an exploratory study therefore no matching of cases to controls will be conducted. Consecutive sampling will be used.

**S2 Table 2: Studies on immune molecule correlation to severe malaria, and sample size estimation for power 0.80 for various biomolecules**

| Variable | Sample size | Effect size | Sample size for  Power 0.80 |
| --- | --- | --- | --- |
| IL 18 [6] | UM-47, SM-43, CM-6 | 1169.7 ± 285.0 in UM versus 2580.0 -4- 2407.0 in SM | Total=52  26 per group |
| IL 4 [7] |  | UM had highest record of IL-4 level followed by CM group and then severe malaria anaemia group (SMA) (255.8±54.13, 102.7±34.88 and 90.95±20.90 pg/ml respectively, P>0.0001) | Total=6  3 per group |
| IL-10 [8] | 14 CM, 11 SM, and 20 UM | mean IL-10 levels of 2812 CM, 2882 SM and 913 pg/ml UM, while 98% of healthy individuals had undetectable (less than 100 pg/ml | Total=12,  6 per group |
| Interleukin 12 [9] | SM 278, UM 249.  Malian children | SM GM 48.9± 54 versus GM 33.8± 7.8 in UM | Total= 210  105 per arm |
| Soluble TNF-α [10] | Gambian children: 178 UM vs non-malaria illness vs 110 CM | Geometric mean TNF in non-fatal CM =51(36-72) and269(170-431) in fatal CM versus24 (20-29) in mild malaria, p<0.05 | Total = 18  9 per group |
| TGF- β [11] | Included 66 children with CM from Tanzania, 28 of whom were sampled | 17.9 ± 1.5 in CM versus 32.6 ± 2.5 in uncomplicated malaria though included adults in control grp, p<0.05 | Total = 4  2 per group |
| Angiopoietin 1 [12] | 67 UM and 69 CM Ugandan children | UM median 25.0 (0.39-64.9)  CM median 9.0 (0.39- 37.5) | Total=194  97 per group |
| Legend: CM- Cerebral malaria, SM- Severe malaria, UM- Uncomplicated malaria, ICAM-1- Intercellular adhesion molecule-1, IL- Interleukin, TNF- Tumor necrosis factor, IFN- Interferon, MAKI- Malaria-associated acute kidney injury. | | | |

**Objective 3**

To verify the power that use of such a sample size would endow the study, the online University of Michigan Genetic Association Study (GAS) Power Calculator was used. Assumptions for the power calculation are based on recommendations by Politi et al.[12] An Alpha of 5% (0.05) was used and since the outcome variable is binary (limited values), the prevalence of MAKI in children with malaria (40%) was used. Additionally, the effect size of genes on complex diseases usually lies in the order of odds ratio 1.1 to 1.5, so an odds ratio of 1.3 was used (12,13) Furthermore, the SNPs were assumed to be common variants (MAF > 5.[12] We assumed a Pearson correlation coefficient for the linkage disequilibrium of r2 = >0.8 for the causal variant and marker SNP.(12)A dominant model for genetic inheritance was assumed i.e AA + Aa vs. aa.[13] Based on these parameters our sample size of 380 has a mere power of 60% thus only descriptive statistics will be used for this section.

Objective 4

Sample size for the RAI study was determined using the Cochrane formula:

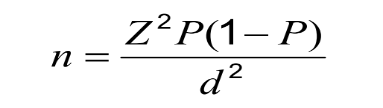

Where n = the required sample size

Z = Normal Standard Deviation taken with a 95% Confidence Interval; set at 1.96.

d = margin of error (0.05)

p = prevalence of AKI in malaria

Assumptions made were a significance level of 0.05, a prevalence of AKI in children admitted with malaria of 0.30 and power of 80%.

Allowing for a 10% loss of data a total sample size of 180 children with malaria and no AKI at admission to the hospital, will need to be recruited into the RAI study.
